# Evaluating the Impact of VA’s Contract Buyout Program: An Analysis of Rural Workforce Recruitment Challenges

**DOI:** 10.64898/2026.02.11.26346089

**Authors:** Aisha Khan, Shannon Kenyon, Patrick O’Mahen, Vivian R. Spencer, Richard SoRelle, Sylvia J. Hysong

## Abstract

**Background:** Approximately 33% of U.S. Veterans live in rural areas, often facing significant barriers to accessing healthcare due to staffing shortages at VA facilities. The Contract Buyout (CBO) program, authorized under the PACT Act of 2022, was designed to address rural healthcare staffing shortages by enabling Veterans Health Administration (VHA) facilities to buy out existing service contracts to work in rural VA facilities. Despite its potential, uptake of the program has been limited, with just 18 hires and $1.5M in expenditures, despite a congressional spending authorization of up to $40M. This evaluation explores the barriers and facilitators in implementation of the CBO program across rural VA facilities.

**Methods:** Using the RE-AIM framework, we conducted a mix-method qualitative evaluation. Semi-structured interviews were completed by 15 interviewees across 8 rural VA facilities, including hiring leaders and physicians. Data were analyzed using rapid qualitative analysis, supported by a descriptive survey to capture the CBO program awareness and experience. We conducted 15 interviews across 8 rural VA medical centers with facility-level hiring leaders and clinicians who were familiar with or involved in using the CBO program.

**Results:** HR-related delays and procedural ambiguities disrupted contract execution and undermined the CBO program’s effectiveness globally. However, sites with strong internal champions and proactive HR teams reported greater success. Interviewees reported the CBO program as a promising tool, though its lack of dedicated funding and resource dissemination hindered broader adoption.

**Conclusion:** The CBO program holds potential as a flexible rural recruitment incentive but faces structural barriers that limit its reach and adoption. Future evaluations should Evaluate whether individual rural VA sites have budgetary flexibility, funding mechanisms, and related resources required to effectively utilize the CBO program.

## Background

Approximately 33% of U.S. Veterans live in rural areas^1^, where persistent staffing shortages at rural Veterans Health Administration (VHA) facilities create significant barriers to timely and essential healthcare access.^2,3^ These rural shortages have been particularly acute among physicians, nurse practitioners, physician assistants and certified registered nurse anesthetists.^2,3^ To address them, Section 902 of the Promise to Address Comprehensive Toxics (PACT) Act of 2022, established the Contract Buyout Program (CBOP; hereafter, referred to as the CBO program), to incentivize healthcare professionals to work in rural VHA facilities.^4^

The CBO program authorizes rural facility leaders to use local funds to buy out existing employment contracts for physicians, nurse practitioners, physician assistants and certified registered nurse anesthetists.^4^ In exchange, medical professionals commit to working at a rural VA facility for four years, accelerating VA’s recruitment and reducing reliance on temporary or community-based providers.^2^

Due to resource limitations, Department of Veterans Affairs’ (VA) Workforce and Management and Consulting (WMC) soft launched the program to accommodate modest initial levels of organic interest and allow for gradual growth. Rather than conduct a large, centralized outreach program, WMC designated one program manager to develop the program’s outreach materials and nurture connections with leadership at rural VA Medical Centers (VAMCs).^5^ Thus far, the program’s impact has been limited. Although the CBO program was authorized for up to $40 Million in potential spending across rural VA facilities^5^, this ceiling reflected an upper limit rather than a direct appropriation. Individual facilities were expected to allocate their own local funds for participation.

In the three years since the program’s inception, rural facilities collectively spent a total of $1.5 million (P. Gaffney, personal communication, April 21, 2025) to support only 18 clinician hires. Despite the program’s potential, VA officials have identified several implementation barriers, including complex administrative processes, insufficient candidate awareness, and limited rural facility support. These challenges have constrained the program’s overall effectiveness, potentially undermining its intended goal of attracting clinicians to rural areas.^5,6^ Here, we present findings from a rapid evaluation of the CBO program’s early-stage implementation, highlighting how the program was understood, adopted, and executed across rural VA facilities.

## Methods

### Design

We conducted a mixed-methods program evaluation of the CBO program, guided by the RE-AIM framework (Reach, Effectiveness, Adoption, Implementation, and Maintenance), as it is the gold standard for assessing implementation impact.^7^ We employed RE-AIM to assess the CBO program’s impact in addressing rural physician shortages and glean insights into contextual and operational factors that could influence each of the framework’s dimensions.

Our evaluation included three components: 1) archival analysis of operational documents and email communications, 2) a brief online survey, and 3) semi-structured interviews with VA staff and leaders involved in rural recruitment and program implementation.^8^ These methods were used to explore target users’ awareness of the CBO program, experiences with its use, perceived barriers, and recommendations for improvement.

### Eliciting Site Context

To inform evaluation design, we reviewed CBO program-related materials provided by WMC such as internal guidance documents, outreach materials, and email communications with regional networks (Vertically Integrated Service Networks, or VISNs) and facilities. These emails detailed which sites were contacted, what information was shared, and how facility-level administration responded. Together, these resources offered insights into how the program was introduced and received across VA sites, including factors such as facility eligibility, geographic distribution, clinical service lines, and specific CBO circumstances.

### Interviewee Recruitment

We identified 33 potential interview candidates in total representing both administrative and clinical roles relevant to CBO program implementation (Figure 1). Eighteen administrators were identified through WMC internal records, and six additional contacts were referred through snowball sampling based on prior CBO program experience. Clinician candidates (n = 9) were identified through WMC documentation. Table 1 lists and defines all interviewee types and **Error! Reference source not found**. summarizes our recruitment strategy and outcomes.

**Table 1.**
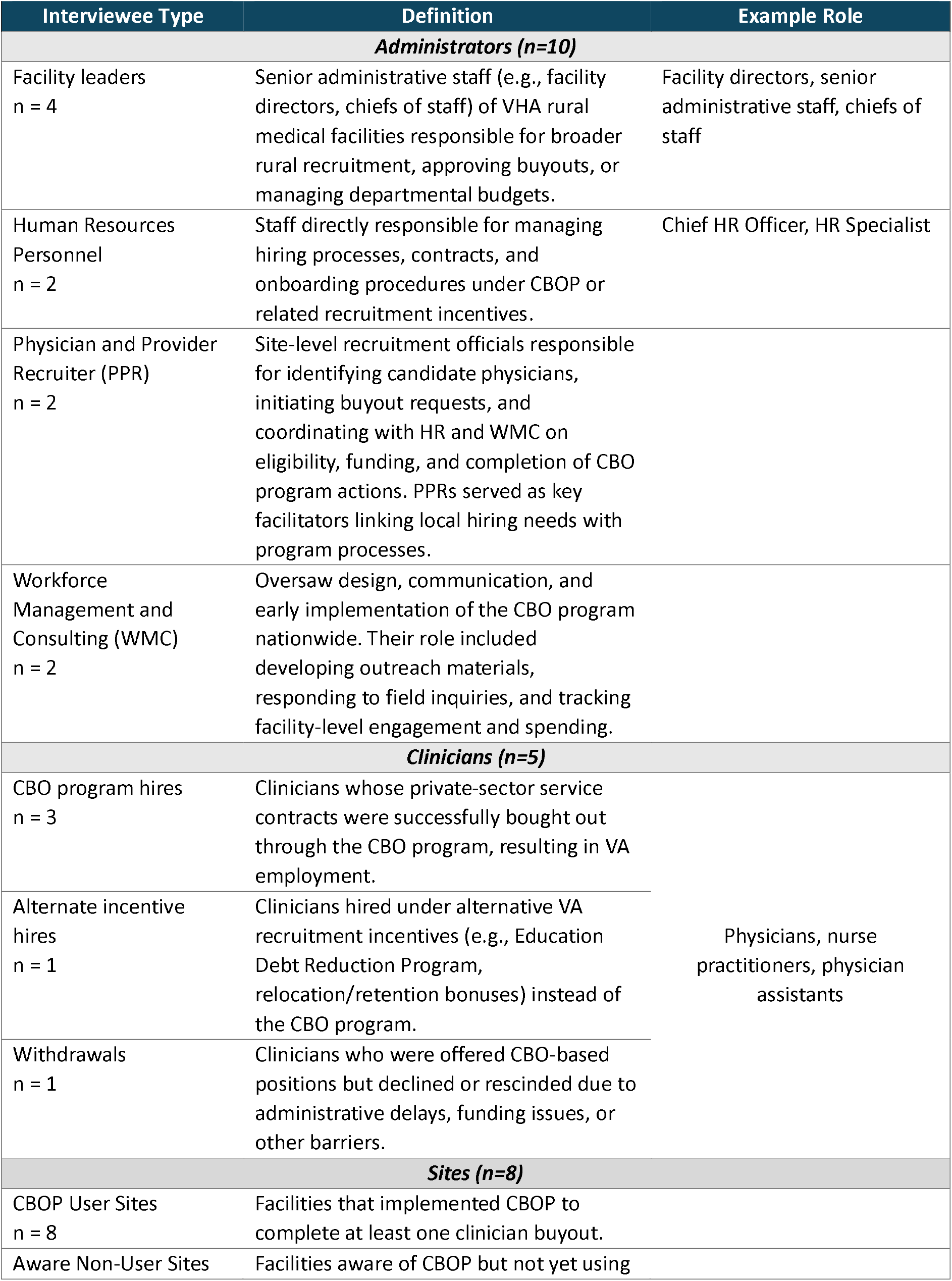
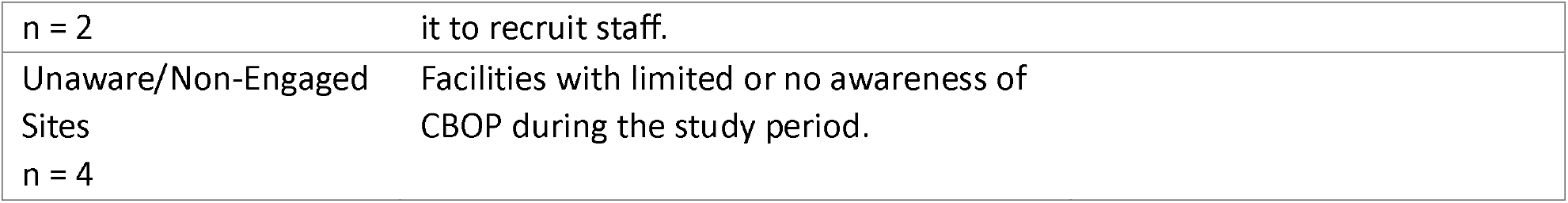
Interviewee Type Nomenclature and Definitions.

**Figure 1.**
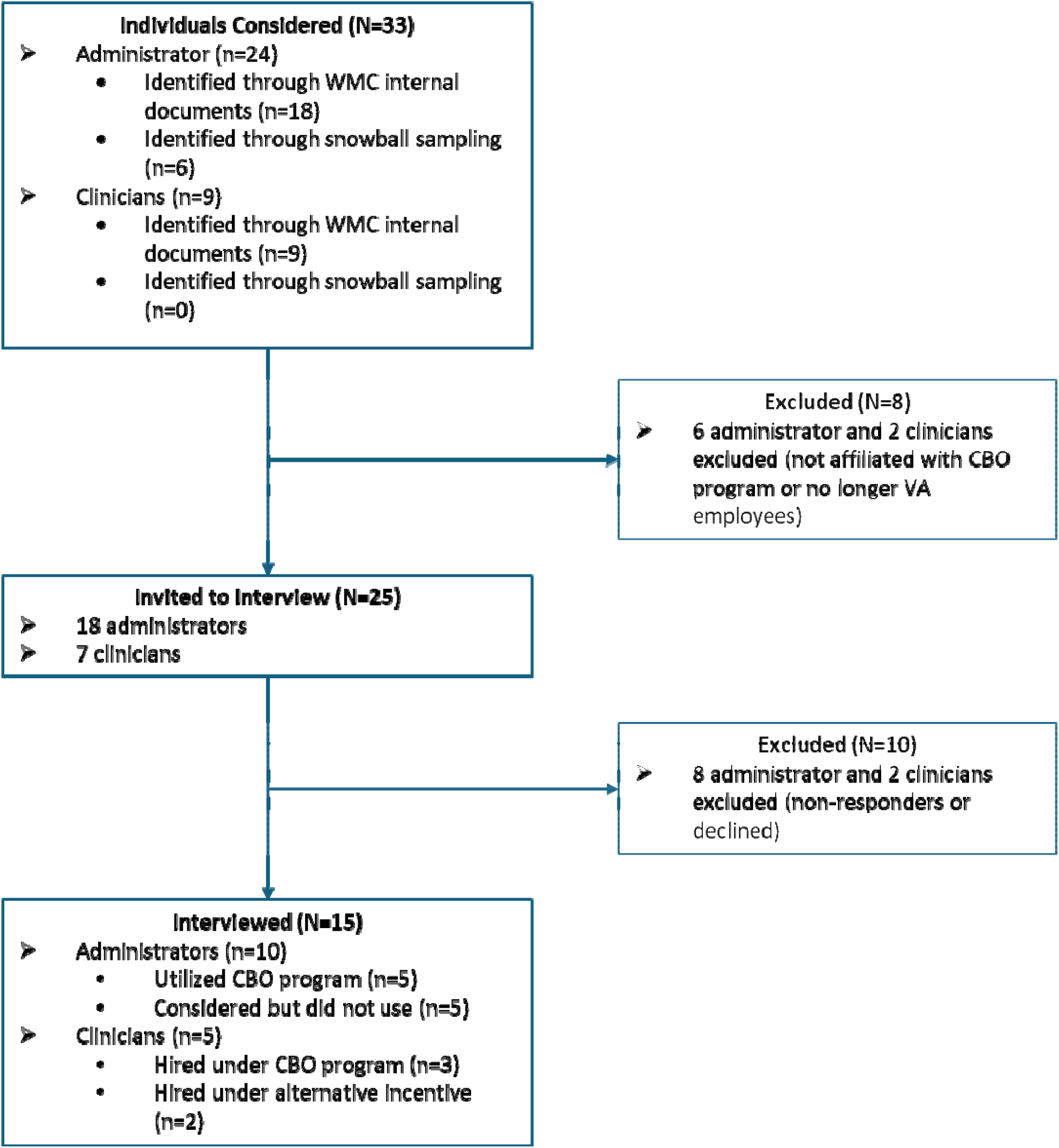
Interviewee Recruitment Flow Diagram.

Of these 33 individuals, 25 were invited to participate. Eight were excluded because they were no longer VA-affiliated or had no direct CBO program involvement. Among those invited, ten did not complete interviews (five declined, five did not respond). The final analytic sample included 15 participants: ten administrators—comprising program managers, HR specialists, and facility leaders—and five clinicians, including physicians hired through either the CBO program or alternate VA incentive pathways. This distribution allowed exploration of both implementation and non-implementation contexts across diverse facilities.

### Procedure

#### Online survey

Prior to interviews, candidates completed a brief online survey designed to capture their roles and levels of the CBO program engagement. This survey served four purposes: 1) assist in tailoring interview prompts for respondents 2) provide contextual descriptive data about respondents 3) confirm respondents as either administrative or clinical hires and 4) generate descriptive data on program awareness and opinions (see Table 1). WMC reviewed the survey to ensure relevance and effectiveness.

#### Interviews

We developed an interview guide using five domains of the RE-AIM framework. Two qualitative evaluators conducted semi-structured interviews from June to August 2025. Interview questions focused on respondents’ awareness of the CBO program, perceived impact, implementation barriers, and facilitators. Evaluators conducted interviews via secure video conferencing and obtained verbal consent prior to each interview. All interviews were audio-recorded and transcribed via Microsoft Teams.

### Data Analysis

The evaluation team reviewed transcripts against recordings for accuracy and de-identified them prior to analysis. Average interview length was approximately 35 minutes. To efficiently identify major themes, barriers, and facilitators related to program uptake, our team employed rapid qualitative analysis, which allowed us to organize and interpret interview data using both deductive and inductive strategies while balancing analytic rigor and timeliness.^9^

Initial coding was guided by a codebook developed a priori and structured around RE-AIM domains. Two team members independently coded each transcript and added emergent codes as needed. They then met to reconcile differences and synthesize domain-specific themes. From these, the team identified recurrent barriers and facilitators within each domain and compared patterns across roles and facility types for consistency and divergence. All coding and data management were conducted using Atlas.ti, Microsoft Excel, and Word.^10^

## Results

Interviewees were geographically diverse and included both clinicians and administrators from sites with varying Veteran population sizes and program experience. Table 2 summarizes key characteristics of the 15 interviewees and eight VAMCs involved in our evaluation.

**Table 2.** Characteristics of Interviewees and Respective Sites.

| Geographic Region<sup>11</sup> | Interviews per site | Interviewee Types | Male | Female | Veteran Population Served |
| --- | --- | --- | --- | --- | --- |
| Continental | 5 | 2 Clinicians/ 3 Administrators | 3 | 2 | 20,000 |
| Continental | 1 | 1 Clinician | 1 | 0 | 26,000 |
| Continental | 1 | 1 Clinician | 1 | 0 | 10,022 |
| Southeast | 1 | 1 Administrators | 0 | 1 | 38,000 |
| Southeast | 2 | 1 Administrators & 1 Clinician | 2 | 0 | 134,000 |
| Southeast | 2 | 1 Administrators & 1 PPR | 1 | 1 | 75,000 |
| WMC | 2 | 2 Administrators | 1 | 1 | *NA |
| Continental | 1 | 1 PPR | 0 | 1 | 40,126 |

Note. Interviewees were grouped into two primary categories: Administrators (n = 10) and Clinicians (n = 5).
PPR = Physician and Provider Recruiter. \*WMC interviewees are linked to multiple facilities

Findings were organized using the RE-AIM framework. Each domain is presented with a summary of implementation barriers and facilitators identified across sites. Illustrative quotations are included to highlight key themes as outlined in Table 3. **Error! Reference source not found**.

**Table 3.**
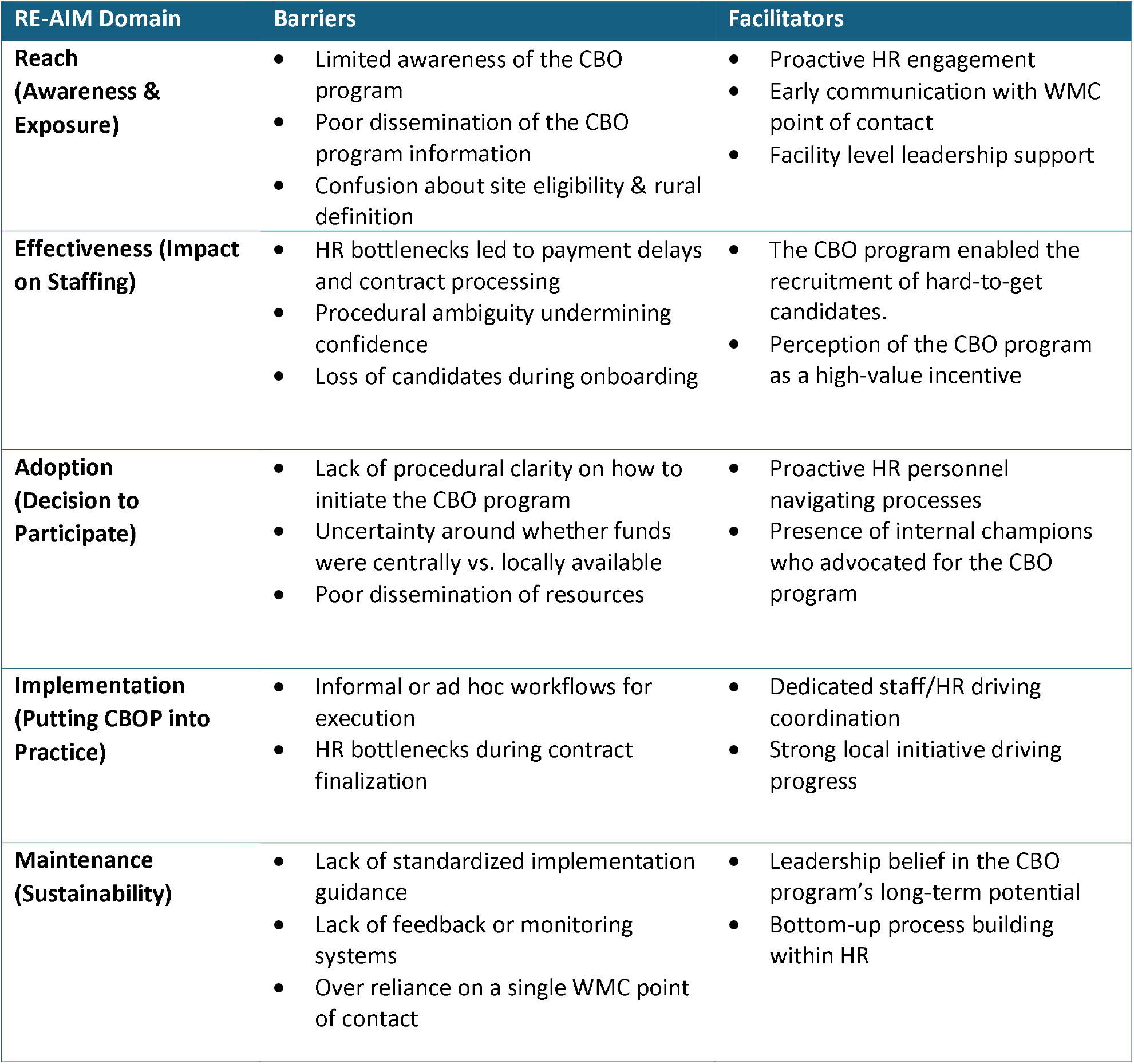
Summary of Barriers and Facilitators by RE-AIM Domain.

## Reach

### Barriers: Limited awareness, inconsistent communication

Interviewees consistently described limited awareness and inconsistent communication about the CBO program across facilities, particularly among clinical leaders and frontline staff. Specifically, nine out of 15 interviewees stated they learned about the program through informal channels or after the opportunity to use it had passed. Four out of 15 interviewees also mentioned confusion about eligibility, including how rural status was determined. For example, one respondent described:

> *“I don’t think it was well known*…*I know some of the providers that came after me*…*actually opted out of [the CBO program] … because no one ever got back to them. So [by] not having information readily available or knowing exactly who to contact made it [difficult]*.*” (Clinician, withdrawal at Non-User Site)*

Narrow visibility hindered early adoption and limited useability even at eligible sites. Additionally, some respondents reported eligibility criteria were poorly understood, leading to missed application opportunities.

### Facilitators: Strong communication, proactive HR involvement

At sites where the CBO program successfully attracted clinicians to VA, respondents credited strong internal communication, proactive HR involvement, and early coordination during the recruitment process. When the program was introduced early in the hiring process, and site leadership understood its requirements, candidates were more likely to access and benefit from it. One CBO program hire shared:

> *“The buyout program allowed me to work at this clinic for a year. I wouldn’t have been able to work here*… *My family lives here, so that would’ve been a big obstacle to live somewhere else for a year. So, [CBOP] made the transition to be very nice and convenient*.*” (Clinician, CBO hiree at CBO User Site)*

## Effectiveness

### Barriers: HR delays & funding mechanism confusion

Although respondents generally viewed the CBO program as an efficient and valuable tool once initiated, its overall effectiveness was often undermined by upstream barriers like HR processing delays and confusion around funding mechanisms, which impacted both applicants’ ability to onboard and sites’ ability to fill needed roles. Notably, 13 out of 15 interviewees expressed discontent with HR onboarding team and a third specifically stated HR delays caused timed-out contacts or clinicians to rescind. Three interviewees recommended developing clear CBO documentation for pre- and post-HR phases of the process to improve transparency. These issues not only disrupted the program’s rollout but also shaped interviewee perceptions of its reliability and feasibility. For instance, interviewees often seemed to misattribute external HR challenges to the CBO program, confusing and frustrating staff. Interviewees described a disconnect between streamlined internal hiring processes at their local facilities and inconsistent practices across VA HR departments when processing the CBO program. A clinician described their experience:

> *“I stumbled across this through interactions with my HR representative from there it gets clunky the process was very time consuming, very incomplete, and led to confusion about what the [CBO] program entailed… Due to the inability to accurately articulate and communicate what the program entailed.” (Clinician, withdrawal at Non-User Site)*

Further confusion, particularly among site administrators, arose regarding the CBO program’s funding mechanism. Five out of 15 interviewees incorrectly assumed that the program would provide external funds, when local sites were expected to allocate their own budgets. This uncertainty sometimes resulted in financial strain or hiring delays. One administrator detailed the frustration of discovering outside funding was not forthcoming:

> *“…our understanding was we could use it [external funding via the CBO program] anywhere and everywhere we needed to use it based upon the discretion of the hospital leadership. With understanding that the money you know we had to justify the dollars that we would spend on a buyout…We found out it was much more restrictive.” (Administrator, facility leader at Non-User Site)*

### Facilitators: Bypassed contractual barriers to clinician recruitment

Despite these barriers, when used effectively, perceived effectiveness was mixed but generally positive. Four facility level administrators explicitly described the CBO program as “worth the effort”, and two favorably compared it to CHOICE reimbursements, viewing the CBO program as a more sustainable solution. Additionally, the program allowed sites to bypass common bottlenecks, such as legal contract delays. As a result, CBO program proved to be a viable mechanism to remove financial penalties tied to contract obligations and recruit clinicians whom administrators might not otherwise consider pursuing. In one case, the CBO program helped a team meet buyout demands of a critical care provider without penalty:

> *“…during that time we had a [an eligible candidate] … he was… very thankful that we worked with him, … through the process and was grateful that was something that we were able to offer him. Otherwise…it wouldn’t have been an option for him to come here because he was working at a facility that was a mile down the road.” (Administrator, facility leader at User Site)*

## Adoption

### Barriers: Burdensome compared to other incentives

Two facility level recruiters described the CBO program as more complex and administratively burdensome than other recruitment tools. Unlike the 3 R’s (Recruitment, Relocation and Retention) incentives, which were described as straightforward and low burden, the CBO program was seen as high effort particularly for physician recruiters.^11^ As one PPR explained:

> *“So typically we make the three R’s… really easy on the doc… I think the [CBO] program probably has additional steps…so there ends up being more of a disconnect and [less willingness] to utilize anything that takes more of their time.” (Administrator, PPR at Aware Non-User Site)*

### Facilitators: Increased awareness & proactive HR

Site administrators that adopted the CBO program described several factors that supported uptake. In some cases, communication through VISN channels, including Chief HR officer calls and program presentations helped increase awareness and understanding of how the program could be used. A facility level administrator recalled:

> *“Through the VISN channels we have Chief HR officer calls … we had them come and do a presentation for our VISN about the program, so a lot of different ways [information was shared].” (Administrator, facility leader at CBO User Site)*

In other cases, local HR taking initiative supported the adoption process. When HR personnel were proactive, the program appeared straightforward to recruiting administrators. One facility level administrator noted:

> *“It was just… there’s this new program and our HR people got in touch with someone, and they knew what to do and they did it. And on the provider side, I think it was pretty seamless and I could go and recruit people.” (Administrator, facility leader at CBO User Site)*

## Implementation

### Barriers: HR delays were misinterpreted as faults within the CBO program

Lack of process clarity emerged as a key barrier to implementation. Two-thirds of interviewees (10 of 15) described confusion on the program’s operational details, including rollout structure, service expectations, and inconsistent communication. Only 7 of 15 interviewees recalled the initial CBO program rollout and an even smaller subset (3 of 15) acknowledged a service commitment -- but could not recall its terms. An equal number (3 of 15) stated they did not remember what they signed and lacked a copy of their agreement. Only 3 interviewees reported no issues with the service commitment, implying inconsistent communication and documentation practices. One frontline clinician, unable to use the program, desired a more structured and transparent process:

> *“We need a clear definition of eligibility, inclusion, exclusion criteria, and we need a product that clearly articulated [the CBO program].” (Clinician, withdrawal at Non-User Site)*

Because the CBO program was often experienced through existing HR systems, many interviewees misinterpreted issues and delays stemming from broader HR, as barriers to the program itself. In other words, although the program was intended as an authorization mechanism, it was often perceived as inseparable from HR’s implementation processes. Consequently, issues with HR responsiveness were interpreted as CBO program problems. As one PPR reflected, a promising candidate withdrew their application after prolonged delays:

> *“Yeah, I think that we just realized that … it was… taking eight months just to get him on boarded so I think…he was like, why would I even go through this?” (Administrator, PPR at Aware Non-User Sites)*

### Facilitators: Engaged and proactive HR

In facilities where the CBO program was implemented more effectively, interviewees described HR staff actively driving the process forward. In contrast to delays typically experienced with HR elsewhere, when HR staff were proactive and engaged, the CBO program was implemented efficiently, reducing delays that had previously discouraged candidates. One facility level administrator emphasized the improved onboarding timelines:

> *“So this goes through HR. And [although] HR has the reputation of being slow…this process seemed to be pretty quick and our HR…were excited about engaging in the process. It got done in a reasonable amount of time and I know it’s quick because. We’ve lost providers when we couldn’t get through HR quick enough…I think it was a matter of weeks.” (Administrator, facility leader at CBO User Site)*

## Maintenance

### Barriers: Lack of clear, standardized communication

Interviewees described key challenges to sustaining the CBO program, particularly the lack of clear, standardized communication about how the program works in practice. Individuals with prior CBO program knowledge perceived it as ambiguous. With unclear responsibilities or defined workflows, interviewees felt unprepared to plan future use or guide others through the program. As one clinician explained:

> *“[I] don’t understand the … minutiae of how the program works… If it’s a situation where I get a candidate and they need to speak with [the CBO program Point of Contact] and facilitate their contract… I don’t…understand who does them or how they’re done. So I guess more communication on how that process works could be helpful on the front end.” (Clinician, withdrawal at Non-User Site)*

Along with communication gaps, there were no clear systems for tracking program use, documenting best practices, or maintaining institutional knowledge over time. Without a shared understanding of how the CBO program should function or who was responsible for each step, interviewees felt long-term program use would prove challenging.

> *“I don’t think [the CBO program] is well known. I know some of the providers that [applied] after me didn’t go through [with it] because no one ever got back to them… and information wasn’t readily available … As a supervisor, I ask myself…, how do I get this person onboarded? What’s the process? Because if they ask me questions, I don’t know [how to answer] it.” (Clinician, withdrawal at Non-User Site)*

### Facilitators: Incentive alignment with rural hiring priorities

Despite concerns about long-term sustainability, those who had used the CBO program emphasized its immediate value in attracting and retaining highly qualified candidates. In some cases, the program made the difference between a candidate accepting or rejecting a VA offer. As one clinician shared how the CBO program aligned with their financial situation:

> *“I asked about [the CBO program] because I had a significant amount [still owed to] my current employer. The idea of having [that] paid off… was appealing because I wouldn’t have to pay that remainder back… and assist me with moving forward with my career.” (Clinician, CBO Hiree at CBO User Site)*

Interviewees (11 of 15) described the CBO program as a valuable tool for rural recruitment and more impactful than traditional incentives because it reshaped how hiring bonuses were perceived and used. Rather than being absorbed into salary budgets or viewed as one-time perks, the CBO program was framed as a strategic investment implying deeper institutional commitment. Both administrators and clinicians emphasized that the CBO program offered a more meaningful and lasting recruitment advantage compared to short-term incentives like the 3 R’s or Education Debt Reduction Program (EDRP), creating a stronger sense of institutional commitment. As one administrator explained:

> *“The 3 R’s … and the EDRP are very important. … But here’s the difference: you’re buying opportunity. I can give someone a recruitment incentive and that is an incentive up until their first paycheck, and then that’s just part of their budget. But what the buyout program does is give me the opportunity to get people. It’s a little more powerful because …everybody wants to make more money, but that impact [fades] after one paycheck. This is different.” (Administrator, facility leader at CBO User Site)*

## Discussion

This evaluation provides one of the first systematic evaluations of the CBO program, offering timely insights into how a national recruitment initiative unfolds across diverse VA facilities. The use of the RE-AIM framework enabled comprehensive analysis, capturing not just uptake and outcomes but also contextual and procedural factors that shaped implementation. Finally, we incorporated diverse perspectives from HR personnel, service line leaders, and facility administrators to further strengthen the validity and nuance of the results.

Interviews with 15 stakeholders at eight rural VAMCs offered clues to the limited current uptake of the VA’s CBO program designed to accelerate rural hiring after the PACT Act. Using the RE-AIM framework to classify the barriers and facilitators to implementing the program, we identified several common themes.

### Lack of information and bureaucratic processes impeded CBO program uptake

The primary hinderances stemmed from unclear information among facility-level administrators and physicians navigating the hiring process. Multiple interviewees (10 of 15) reported confusion about the CBO program eligibility and financing. About a third of respondents (4 of 15) noted they were unsure of whether their VAMC met the criteria for program inclusion, impeding uptake. In total, 13 interviewees expressed dissatisfaction with the HR onboarding process, with some unaware that the program lacked a centralized funding mechanism and instead required local sites to fund their own CBO. This confusion led to delayed budget approvals and, sometimes, the loss of candidates when local funding was unavailable. Sparse national guidance reinforced the dearth of sites’ knowledge and led to questions of program sustainability.

However, some information problems were beyond program implementation. The slow VA hiring process and unclear HR procedures led to losing several promising candidates. Interviewees noted greater HR awareness of the CBO program and more flexibility in its application could have resulted in additional hires.

### Local champions implemented the CBO program effectively

Despite these barriers, the CBO program did experience patterns of success among several VAs with common features. Specifically, four area level administrators described instances where the program helped them retain strong candidates who would have otherwise left. In these cases, clear internal communication and proactive coordination between HR and clinical leadership were key. Thus, once a VA achieved an initial hiring success, repeating the process became easier: two of the 15 eligible facilities accounted for 40% of total CBO program hires.

### Existence of hiring incentives is not enough to guarantee effective use

Past evaluations of VA’s rural recruiting efforts suggested that mere existence of hiring incentives is insufficient to attract clinical staff unless hiring administrators can effectively deploy them in the recruiting process.^12^ Although our interviews showed circumstances under which the CBO program could be effective, questions remain about its effectiveness compared to other VA hiring initiatives, notably the 3 Rs. Our evidence is mixed, with several interviewees noting it was easier to employ the 3 Rs as an effective hiring tool, versus one who enthusiastically supported the CBO program with its unique appeal and reach.

WMC has suggested increasing resources supplied to internal partnering, boosting collaboration with VISNs and local centers to increase knowledge of the CBO program and its integration with other incentives. The CBO program was also identified as a potentially effective tool for urban recruitment, alongside rural, due to greater market competitiveness in urban areas. However, implementing these strategies would require additional dedicated staffing, as the initiative is currently supported by only one individual.

Finally, it is unclear whether simply adding more hiring incentives increases VA hiring capacity. Associated bureaucratic procedures limit effectiveness of any given incentives. VA HR professionals face considerable bureaucratic challenges in hiring processes; clear information on existing hiring incentives prevents bottlenecks for effective use. However, requiring knowledge on more incentives perversely increases their administrative burden.

By statute, the CBO program targets rural areas. However, because the program lacks a dedicated funding source to finance its use, constrained budgets at rural VAs limit effective deployment. Supporting the CBO program with a proportional designated facility allowance could augment attractiveness of the program to rural administrators and improve its practical effectiveness.

### Which incentives work best for rural areas?

Although our interviews showed circumstances under which the CBO program could be effective, large questions remain about its relative effectiveness against other extant VA hiring initiatives. For example, is a contract buy-out more effective than signing bonuses? Nor is it clear that adding more hiring incentives betters VA hiring when associated bureaucratic procedures hinder any incentive.

## Limitations

While the sample included a diverse mix of facility-level administrators, PPR, and clinicians, the perspectives may not represent all stakeholder roles or facility types. Because the CBO program was in early stages of rollout during data collection, interviewee feedback likely reflects initial implementation challenges, which may not represent long-term outcomes. Additionally, interviewees had variable levels of familiarity with the program, potentially impacting their interpretation of its value or feasibility. Although efforts were made to include both users and non-users of the program, selection bias is still likely: for example, we could not interview unhired clinicians may but have been if they had known the CBO program was available.

## Conclusion

Despite its limited early scope, the CBO program represents a promising strategy to address long-standing recruitment challenges in competitive labor markets. Although well-intended in principle, the program’s early implementation revealed systemic barriers limited awareness, disrupted execution, and hindered sustainability. Improving information sharing about the CBO program both with recruiters and HR teams and providing a program designated funding source may increase its usefulness and utilization, though further evaluation is needed to determine whether it plays a unique role in recruitment or is merely a redundant incentive.

## Data Availability

The data that support the findings of this project are available upon written request from the corresponding author. The data are not publicly available, per VA privacy policies.

